# After the vaccine era: sequencing platform investments as the childhood pneumonia spectrum diversifies

**DOI:** 10.64898/2026.09.01.26361934

**Authors:** Deze Li, Hao Chen, Xiaotong Wang, Chen Shen

## Abstract

**Background:** The historic decline in childhood pneumonia mortality was driven substantially by single-pathogen vaccines against *Haemophilus influenzae* type b (Hib) and *Streptococcus pneumoniae*. Yet the pathogen spectrum underlying child pneumonia deaths is diversifying: the effective number of pathogens rose from 5.57 in 1990 to 9.94 in 2023, and the residual burden is shifting toward opportunistic and hospital-associated pathogens for which no licensed childhood vaccines exist. This paper asks how resources should be sequenced between single-pathogen interventions and platform investments as this transition proceeds.

**Methods:** We analyzed Global Burden of Disease Study 2023 deaths from 29 pathogens in ages 0–19 years by super-region, combined with WHO/UNICEF Estimates of National Immunization Coverage (WUENIC) for PCV3 and Hib3. We quantified the spectrum transition under two denominators (26- and 29-pathogen calibers), constructed a share-by-intervenability matrix assigning each pathogen to a dominant intervention channel (vaccine-reachable, mixed, platform-sensitive) under explicit classification rules, compared platform-sensitive deaths with a transparently computed scenario of residual vaccine-preventable deaths, and cross-classified pathogens by age tropism and poverty lock. We anchored platform interventions to verified published evidence.

**Results:** The vaccine-preventable group share fell from 54.0% to 40.2% while the opportunistic/hospital group rose from 18.1% to 23.1% (29-pathogen caliber, 1990–2023). Super-region vaccine coverage showed no significant association with pathogen-share change (PCV3 Spearman ρ = 0.108, p = 0.818; Hib3 ρ = −0.036, p = 0.939), a null result we report as evidence that simple coverage–burden correlations do not hold at the regional level, not as evidence against vaccine value. In 2023, vaccine-reachable pathogens accounted for 441,410 deaths (45.7%, channel including COVID-19), mixed for 126,926 (13.1%), and platform-sensitive pathogens for 396,995 (41.1%). Platform-sensitive deaths were 2.9–5.1 times the scenario estimate of residual vaccine- preventable deaths (52,435–77,512). Nine of 14 classifiable pathogens fell into the poverty-locked, infant-tropic cell (480,922 deaths; Fisher OR = 9.0, p = 0.1758).

**Conclusions:** The marginal value of single-pathogen strategies declines as the spectrum diversifies and residual deaths concentrate in platform-sensitive, poverty-locked, infant-tropic pathogens. Vaccine scale-up remains a certain and sizeable opportunity; the next increment of marginal resources should increasingly fund platform capabilities (oxygen systems, antimicrobial access and stewardship, infection prevention and control, referral, and nutrition) delivered as a package to the populations where the residual burden is locked.

## 1. Introduction

Few global-health achievements rival the decline in childhood pneumonia mortality over the past three decades. Pneumonia remains the leading infectious cause of death in children and a dominant component of the unfinished communicable-disease agenda in the young, but annual deaths in ages 0–19 years have fallen steeply since 1990 ^1–3^. A large share of that decline is attributable to a single-pathogen strategy: the introduction and scale-up of vaccines against two bacterial pathogens, *Haemophilus influenzae* type b (Hib) and *Streptococcus pneumoniae* (pneumococcus), delivered through Hib-containing and pneumococcal conjugate vaccines (PCV) and supported by donor financing, accelerated introduction programs, and market-shaping instruments designed around individual products ^2,3^. The strategy was coherent with the epidemiology of the time. In 1990, a single pathogen, pneumococcus, accounted for 37.9% of all pathogen-attributed deaths in ages 0–19 years (recomputed here from GBD 2023 estimates; Methods), and a small set of vaccine-preventable pathogens dominated the mortality spectrum. Single-pathogen tools aimed at a concentrated target produced concentrated returns, and the policy apparatus of the pneumonia community (targets, financing, monitoring indicators) grew up around that logic.

The target, however, has been moving. Global immunization coverage has expanded dramatically: WUENIC estimates place global PCV3 coverage at 4% in 2008 and 66% in 2023, and Hib3 coverage at 77% in 2023, leaving global uncovered fractions of 34% and 23% respectively ^4^. In parallel, the composition of the residual burden has diversified. Our analysis of Global Burden of Disease Study 2023 (GBD 2023) estimates across 29 pathogen nodes shows the vaccine-preventable group share falling from 54.0% to 40.2% between 1990 and 2023, while a group of five opportunistic and hospital-associated pathogens (*Pseudomonas aeruginosa*, *Staphylococcus aureus*, non-tuberculous mycobacteria, *Acinetobacter baumannii*, *Klebsiella pneumoniae*) rose from 18.1% to 23.1%. The effective number of pathogens (the reciprocal of the Herfindahl– Hirschman index, equivalent to the inverse-Simpson diversity measure or Hill number of order 2 ^5^) rose from 5.57 to 9.94 over the same period. By this measure the mortality spectrum has shifted from a single-killer regime toward a multi-pathogen equilibrium in which no single organism dominates the way pneumococcus did in 1990.

This transition changes the economics of intervention choice. When a small number of vaccine-preventable pathogens dominate, the marginal value of vaccine investment is high and approximately linear in coverage: each additional covered birth cohort removes a predictable fraction of a large, well-identified pool. As the spectrum diversifies, three things happen. First, each additional single-pathogen tool addresses a smaller slice of the total, and the pipeline of licensable childhood vaccines thins as the remaining large-share pathogens are exhausted. Second, the residual burden increasingly comprises pathogens for which no licensed childhood vaccine exists and none is imminent. These deaths are compressible not by immunization but by platform interventions that act across pathogens on the shared severe-disease pathway: oxygen systems for hypoxaemia, a common and long-neglected complication of childhood pneumonia and a recognized risk factor for death ^6,7^; timely access to appropriate antimicrobials, paired with stewardship; infection prevention and control; severe-case referral and grading; and nutrition. Third, the residual burden becomes more geographically concentrated (in our data, roughly four-fifths of deaths in every intervention channel occur in the sub-Saharan Africa and South Asia super-regions), raising the value of regionally delivered platform capability relative to globally uniform products.

Against this background, the strategic question for child-pneumonia resource allocation is no longer only “how fast can we close vaccine coverage gaps?” but “in what sequence should marginal resources move from single-pathogen interventions toward platform investments?” The question is comparative, not adversarial: vaccine coverage gaps remain a real and quantified opportunity, and any credible sequence must finish that agenda rather than abandon it. Nor is the concept of platform care new: integrated case management and facility strengthening have been part of child-health policy for decades, but platform components have typically been funded as residual line items behind product-specific verticals, justified by syndrome-level arguments rather than by a quantified account of how much of the remaining pathogen spectrum only they can reach. Scott and English anticipated in 2008 that successful Hib and pneumococcal vaccination would diversify the causes of childhood pneumonia and render diagnosis and management built around two dominant organisms obsolete ^8^; the integrated Global Action Plan for Pneumonia and Diarrhoea (GAPPD) codified a protect–prevent–treat package, including oxygen, as joint WHO/UNICEF policy ^9^; and the Every Breath Counts coalition now advocates the same integrated agenda ^10^. What the diversifying spectrum changes is the size of that residual claim. The relative scale, location, and age profile of the remaining pools matter for what should be funded next, and to our knowledge those quantities have not been placed side by side in a single quantitative framework; the increment of this paper over that lineage is measurement rather than concept—a 29-pathogen quantification of the spectrum transition, an explicit share-by-intervenability classification, and a transparent scenario comparison that together turn the integrated-strategy argument into a sequencing tool.

This paper provides that framework. Using GBD 2023 deaths for 29 pathogens in ages 0–19 years, we (i) quantify the spectrum transition under two pathogen denominators; (ii) classify all 29 pathogens into a share-by-intervenability matrix that locates each pathogen’s dominant intervention channel (vaccine-reachable, mixed, or platform-sensitive) under explicit, rule-based criteria; (iii) compare the measured scale of platform-sensitive deaths with a transparently specified scenario of residual vaccine-preventable deaths; and (iv) overlay age tropism and poverty lock to identify where a combined vaccine-plus-platform package should be concentrated. We anchor the platform side of the argument to verified published evidence on oxygen systems and hypoxaemia care. We compute no costs in this study; all cost and effect figures cited from the literature are identified as published values carrying their original certainty ratings.

## 2. Methods

### 2.1 Data sources

Pathogen-specific death estimates were taken from GBD 2023, which provides cause-of-death estimates for 292 causes in 204 countries and territories, 1990–2023 ^1^. We used 29 pathogen nodes attributable to lower respiratory infection and related syndromes in ages 0–19 years, aggregated globally and by GBD super-region; etiological attribution follows the GBD lower respiratory infection framework, in which deaths are redistributed across etiologies under a counterfactual model ^2,3^. GBD estimates are modeled quantities; we use point estimates throughout and return to this limitation in the Discussion. Two denominators are used and declared once here: the 29-pathogen caliber (the full node set, used as the primary caliber in the Results) and the 26-pathogen caliber (a legacy denominator excluding COVID-19 and two recently added nodes, retained for comparability with earlier analyses and to show that the transition is not an artifact of node-set expansion). Where the two calibers yield different figures, both are reported in Table 1; elsewhere the 29-pathogen caliber is used. Poverty concentration is expressed as each pathogen’s share of deaths occurring in the sub-Saharan Africa and South Asia super-regions (SSA+SA), the two super-regions that carry the large majority of child pneumonia deaths. Immunization coverage was taken from WUENIC global aggregate rows for PCV3 and Hib3 (2025 revision, released 15 July 2026 and retrieved via the WHO Global Health Observatory API in August 2026) ^4^; WUENIC zero values denote non-introduction and are not treated as measured zero coverage. . All tabulations were recomputed from the underlying CSV extracts.

**Table 1.** Spectrum transition, four time points, two calibers (ages 0–19, global).

| **Year** | **Vaccine group share, 29-caliber (%)** | **Opportunistic/hospital share, 29-caliber (%)** | **Vaccine trio share, 26-caliber (%)** | **Opportunistic share, 26-caliber (%)** | **effN (29-node)** |
|---|---|---|---|---|---|
| 1990 | 54.0 | 18.1 | 54.6 | 22.9* | 5.57 |
| 2010 | 51.9 | 18.8 | 52.0 | 24.0 | 6.13 |
| 2019 | 44.9 | 22.1 | 41.9 | 28.2 | 8.94 |
| 2023 | 40.2 (45.7 with COVID-19) | 23.1 | 38.7 | 31.3 | 9.94 |

\* 26-caliber 1990 value is 22.846%, quoted as 22.9% at one decimal place. Vaccine group (29-caliber): pneumococcus + Hib + influenza + pertussis. Opportunistic/hospital group: *Pseudomonas aeruginosa*, *Staphylococcus aureus*, non-tuberculous mycobacteria, *Acinetobacter baumannii*, *Klebsiella pneumoniae*. effN = 1/Σp².

### 2.2 Quantifying the spectrum transition

We defined two pathogen groups. The vaccine-preventable group (hereafter the vaccine group) comprises *Streptococcus pneumoniae*, *Haemophilus influenzae*, influenza, and pertussis (with COVID-19 shown as an alternative 2023 caliber). The opportunistic/hospital group comprises *Pseudomonas aeruginosa*, *Staphylococcus aureus*, non-tuberculous mycobacteria, *Acinetobacter baumannii*, and *Klebsiella pneumoniae*. Group shares were computed as group deaths divided by the yearly all-pathogen total at four time points (1990, 2010, 2019, 2023). Spectrum diversity was summarized as the effective number of pathogens, effN = 1/Σp², where p is each node’s share ^5^.

To test whether coverage expansion maps onto share decline, we computed Spearman rank correlations between super-region median WUENIC coverage (2015, the midpoint of the transition window) and the 2010–2023 change in pathogen share across the seven super-regions, separately for PCV3 (pneumococcal share) and Hib3 (Hib share). We chose the super-region level because WUENIC-derived covariates have already been incorporated into GBD etiological models ^2^; an independent, aggregate cross-check is therefore informative even at n = 7. We report the resulting null associations as-is, with exact p-values, and interpret them as descriptive ecology rather than causal evidence in either direction.

### 2.3 Share-by-intervenability matrix

Each of the 29 pathogens was assigned to a dominant intervention channel under explicit rules: **vaccine-reachable** (a licensed vaccine exists and is in routine immunization programs: pneumococcus, Hib, influenza, pertussis, COVID-19; n = 5); **mixed** (partial or emerging immunization with a dominant delivery-platform component: tuberculosis, where BCG gives partial protection and the diagnostic-treatment platform dominates; respiratory syncytial virus (RSV), where maternal vaccine and long-acting monoclonal antibodies are newly available but supportive oxygen care remains the backbone; and group B streptococcus (GBS), where vaccines are in development and intrapartum antimicrobial prophylaxis is the current tool; n = 3); and **platform-sensitive** (all remaining bacterial, fungal, and mixed-pathogen nodes with no licensed childhood vaccine; n = 21). Channel assignment is a rule-based judgment reflecting the dominant *current* channel, not a claim about future licensability; the per-pathogen rationale is listed in the supplementary matrix, and the sensitivity of channel totals to the mixed assignments is discussed as a limitation. Channels were cross-classified by share tier (high: >50,000 deaths in 2023; medium: 5,000–50,000; low: <5,000) and by SSA+SA concentration, so that each cell can be read as “death scale × dominant channel × poverty concentration.”

### 2.4 Platform-sensitive scale versus residual vaccine-preventable space

Platform-sensitive deaths in 2023 are measured values (sums of GBD point estimates), reported for the five-pathogen opportunistic/hospital group and for the full 21-pathogen platform channel. Residual vaccine-preventable space—more precisely, residual pneumococcal-and-Hib space—is a **scenario**, not a measurement, and its pathogen scope is declared up front: the scenario covers pneumococcus and Hib only, and excludes residual influenza and pertussis deaths (142,760 deaths in 2023), for which no comparable global uncovered fraction is computed here. The computation is fully transparent: low scenario = 227,976.5 (pneumococcal deaths, 2023) × 0.23 (the smaller global uncovered fraction, Hib3, used as a conservative bracket and not as a Hib-based estimate) = 52,435; high scenario = 227,976.5 × 0.34 (PCV3 uncovered fraction) = 77,512; Hib add-on = 17,774.8 × 0.23 = 4,088. Assumptions: deaths are distributed in proportion to coverage; the vaccine is fully effective within reach; indirect effects, serotype replacement, and the higher baseline risk of uncovered populations are ignored. Both the restricted pathogen scope and the higher baseline risk of uncovered populations bias the scenario estimate of the residual downward: the true residual vaccine-preventable space is most plausibly larger than these figures, and platform-to-residual ratios computed against them are correspondingly upper bounds. These figures are not cost-effectiveness conclusions.

### 2.5 Age tropism and poverty lock

For the 14 pathogen nodes with computable age-tropism and lock indices (the remaining nodes lack stable age-split or regional estimates), pathogens were cross-classified by age tropism (infant-tropic, where the share-age index (SAI) < 1 indicates concentration of deaths in infancy; older-tropic, SAI ≥ 1) and poverty lock (SSA+SA share above or below the 80.59% all-spectrum reference, the deaths-weighted average concentration across the spectrum). Cell enrichment was tested with a two-sided Fisher exact test; given only 14 classifiable nodes, the test is underpowered, and we report the exact p-value regardless of the conventional significance threshold.

Robustness of the 2×2 classification was assessed by perturbing both thresholds: the lock reference by ±5 percentage points (75.59% and 85.59%) and the SAI threshold by ±5% relative (0.95 and 1.05), singly and jointly, recomputing cell membership, cell deaths and the Fisher exact test in each scenario. Stability was further assessed with 500 bootstrap resamples of the 14 nodes (seed 42); because set membership is a deterministic function of each node’s two indices, resampling cannot alter any label, so the bootstrap tracked the Fisher odds ratio and p value only.

The full threshold-scenario grid and bootstrap output are provided in Additional file 7. The share tiers (>50,000; 5,000–50,000; <5,000 deaths) are absolute-count cutoffs and the intervention-channel assignment is a non-numeric rule, so neither admits a percentage-point perturbation.

### 2.6 Literature anchors

Platform-intervention effect and cost figures were taken from published studies verified against PubMed records (citations and abstract-level numbers checked individually): a multihospital effectiveness study of improved oxygen systems in Papua New Guinea ^11^; a systematic review, meta-analysis, and cost-effectiveness analysis of oxygen systems strengthening ^12^; systematic reviews of hypoxaemia prevalence in childhood pneumonia and acute lower respiratory infection ^6,7^; and the WHO manual on oxygen therapy for children ^13^. Evidence certainty is reported as rated by the original authors, including low-certainty ratings where assigned. Cost figures are literature values, not computations of this study. Consistent with the declared boundary, the framework is a prioritization tool, not a cost-effectiveness calculation; no costs were computed in this study.

## 3. Results

### 3.1 The spectrum transition

Table 1 and Figure 1 summarize the transition under both calibers. In the primary 29-pathogen caliber, the vaccine group share fell from 54.0% in 1990 to 51.9% in 2010, 44.9% in 2019, and 40.2% in 2023 (45.7% if COVID-19 is added to the vaccine group as an alternative caliber). The decline was modest during the first two decades (2.1 percentage points between 1990 and 2010) and accelerated thereafter (11.7 percentage points between 2010 and 2023), consistent with the delayed but compounding effect of PCV introduction in high-burden countries after 2008 ^4^. The opportunistic/hospital group moved in the opposite direction, rising from 18.1% in 1990 to 18.8% in 2010, 22.1% in 2019, and 23.1% in 2023; its absolute share gain of five percentage points over the full period understates its relative growth of 28%. The 26-pathogen caliber shows the same transition in steeper form (Table 1), confirming that it is a property of the burden itself rather than of the node set chosen to measure it. Spectrum diversity rose monotonically under either caliber: the effective number of pathogens increased from 5.57 in 1990 to 6.13 in 2010, 8.94 in 2019, and 9.94 in 2023—meaning the 2023 spectrum is spread across the equivalent of roughly ten equally sized pathogens, against fewer than six in 1990.

**Figure 1.**
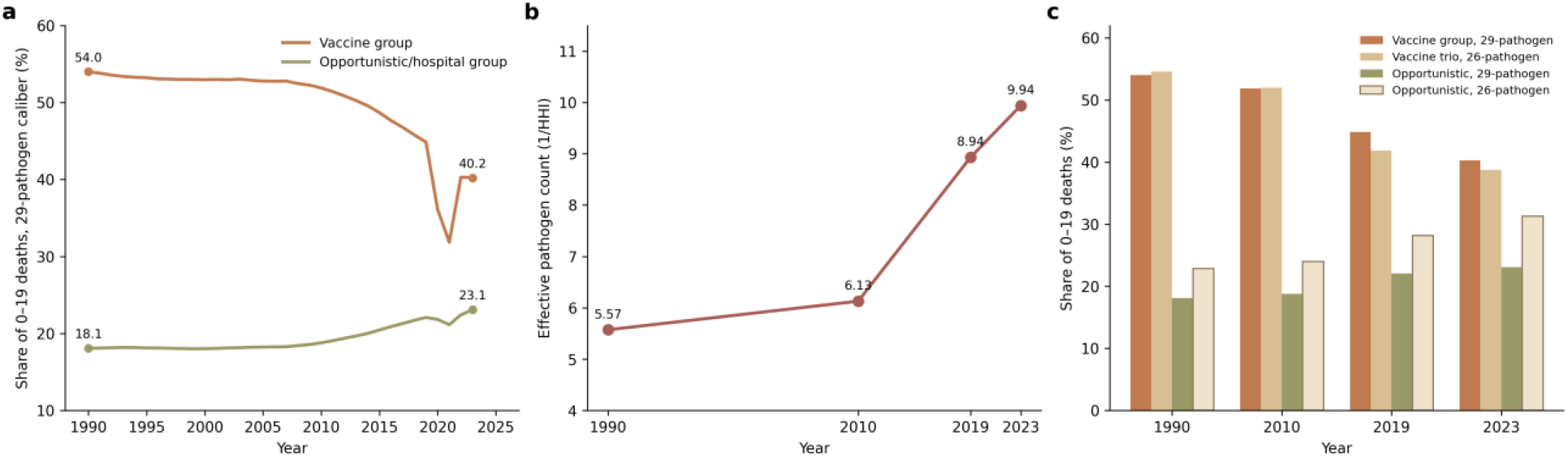
Spectrum transition, 1990–2023. (a) Vaccine-group share (54.0% to 40.2%) and opportunistic/hospital-group share (18.1% to 23.1%) of global deaths at ages 0–19 years, 29-pathogen caliber. (b) Effective pathogen count (1/HHI), 29-pathogen caliber: 5.57 (1990), 6.13 (2010), 8.94 (2019), 9.94 (2023). (c) Dual-caliber comparison at 1990, 2010, 2019 and 2023: vaccine group under the 29-pathogen caliber (54.0/51.9/44.9/40.2%) versus the vaccine trio under the 26-pathogen caliber (54.6/52.0/41.9/38.7%); opportunistic/hospital group under the 29-pathogen caliber (18.1/18.8/22.1/23.1%) versus the 26-pathogen caliber (22.85/24.0/28.2/31.3%).

The dose–response analysis returned a null result, which we report plainly and in full (Figure 5). Across the seven super-regions, median PCV3 coverage was not significantly associated with the 2010–2023 change in pneumococcal share (Spearman ρ = 0.108, p = 0.818, n = 7), and Hib3 coverage was likewise unassociated with the change in Hib share (ρ = −0.036, p = 0.939, n = 7). Pneumococcal share fell in all seven super-regions, so the direction of the vaccine effect is visible; what is absent is any linear dose relationship between the regional coverage gradient and the magnitude of share decline. We interpret this null in a narrow way. It is not evidence against vaccine value: the individual-level efficacy of Hib and pneumococcal conjugate vaccines is established by randomized trials and post-introduction surveillance ^14,15^, and the universal direction of share decline is consistent with it. What the null shows is that a simple coverage– burden correlation does not hold at the regional level. The residual burden, and the pace at which shares transition, appear to be co-driven by structural and platform-side factors (treatment access, case management, nutrition, and risk-factor decline) that vary independently of coverage. Two implications follow. First, the transition cannot be attributed to vaccines alone, and the historical contribution of platform-side investments to pneumonia mortality decline is unlikely to be negligible. Second, the remaining vaccine-sensitive burden will not necessarily fall in proportion to further coverage gains unless those structural factors move as well—an empirical point in favor of pairing the two channels rather than sequencing them in isolation.

### 3.2 The share-by-intervenability matrix in 2023

Table 2 and Figure 2 locate all 29 pathogens in the matrix. In 2023 the five vaccine-reachable pathogens accounted for 441,410 deaths, 45.7% of the spectrum, with 78.6% of those deaths in SSA+SA. The three mixed pathogens accounted for 126,926 deaths (13.1%; 85.1% in SSA+SA), and the 21 platform-sensitive pathogens for 396,995 deaths (41.1%; 81.4% in SSA+SA). The tier breakdown sharpens the picture. Within the vaccine channel, three high-tier pathogens (pneumococcus, pertussis and COVID-19) carried 393,829 deaths and two medium-tier pathogens (influenza and Hib) a further 47,581; there are no low-tier vaccine-reachable pathogens, reflecting that the spectrum’s largest single blocks are still, in principle, vaccine-addressable. The mixed channel is dominated by tuberculosis alone (87,764 deaths, 87.1% in SSA+SA—the highest concentration of any large pathogen in the matrix), with RSV and group B streptococcus in the medium tier. Within the platform channel, two high-share pathogens (*Klebsiella pneumoniae* and *Pseudomonas aeruginosa*) accounted for 137,436 deaths, of which 83.1% occurred in SSA+SA; twelve medium-share pathogens carried a further 243,257 deaths (80.7% in SSA+SA); and seven low-share pathogens accounted for 16,301 deaths.

**Figure 2.**
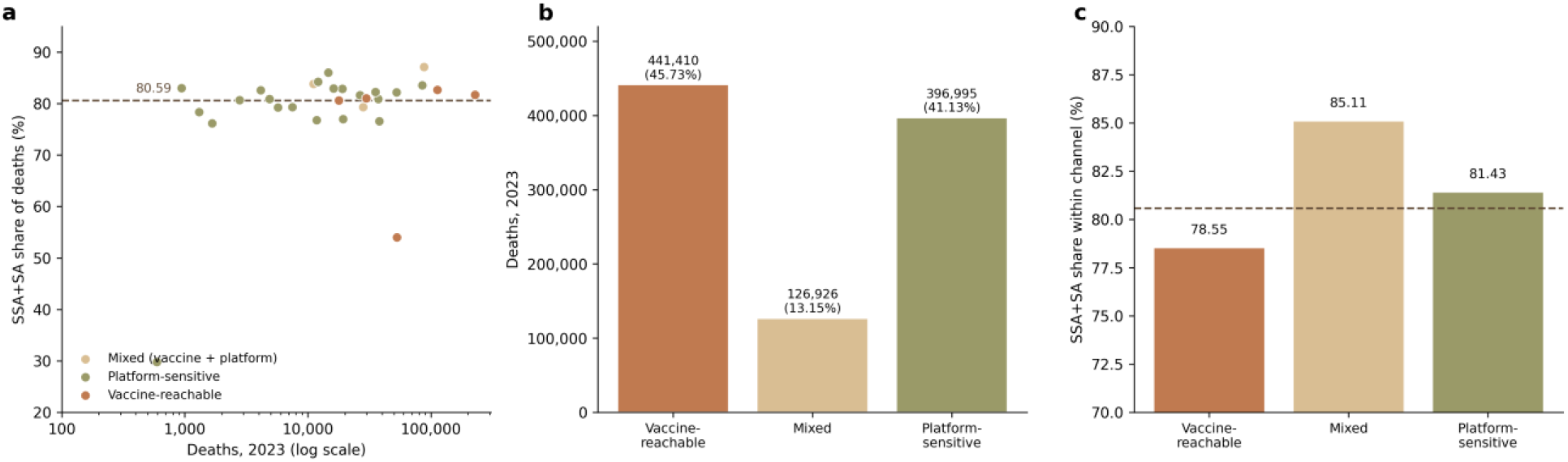
Share-by-intervenability matrix, 2023. (a) Scatter of the 29 pathogens: 2023 deaths (log scale) versus SSA+SA share of deaths, colored by intervention channel; dashed line, 80.59% lock reference. (b) Deaths by channel: vaccine-reachable 441,410 (45.73%), mixed 126,926 (13.15%), platform-sensitive 396,995 (41.13%). (c) SSA+SA share within channel: vaccine-reachable 78.55%, mixed 85.11%, platform-sensitive 81.43%.

**Table 2.** Share-by-intervenability matrix, 2023 (29 pathogens, ages 0–19).

| Channel | Pathogens (n) | Deaths 2023 | Share of spectrum (%) | SSA+SA share (%) | High tier (>50k) | Medium tier (5k–50k) | Low tier (<5k) |
|---|---|---|---|---|---|---|---|
| Vaccine-reachable | 5 | 441,410 | 45.7 | 78.6 | 3 pathogens / 393,829 / 78.3% | 2 / 47,581 / 80.9% | — |
| Mixed (vaccine + platform) | 3 | 126,926 | 13.1 | 85.1 | 1 (tuberculosis) / 87,764 / 87.1% | 2 / 39,161 / 80.6% | — |
| Platform-sensitive | 21 | 396,995 | 41.1 | 81.4 | 2 (*Klebsiella*, *Pseudomonas*) / 137,436 / 83.1% | 12 / 243,257 / 80.7% | 7 / 16,301 / 78.9% |

Tier cells show number of pathogens / deaths / SSA+SA share. Channel assignment is rule-based (per-pathogen rationale in supplement). Tier figures are rounded; channel totals are computed at full precision, so tier sums may differ from channel totals by up to one death. Channel percentage shares (45.7 + 13.1 + 41.1) sum to 99.9% owing to rounding to one decimal place.

Three readings of the matrix follow. First, the platform channel is not a long tail of trivia: its high and medium tiers together (380,693 deaths) approach the size of the entire vaccine channel. Second, the matrix’s first-priority cell for platform investment is the high-share platform-sensitive cell: large in absolute deaths, unaddressed by any licensed vaccine, and more poverty-concentrated (83.1%) than the vaccine channel (78.6%). At country level this cell is tightly concentrated: of its 137,436 deaths in 2023, India (27,657), Nigeria (21,270) and Pakistan (7,938) together account for 41.4% (country-level detail in Additional file 2). Third, poverty concentration is high and uniform across cells (78.3%–87.1%), meaning the geographic delivery target barely depends on which cell is prioritized—every priority cell points at the same two super-regions. This uniformity simplifies the design problem: whatever the channel mix, the delivery geography is SSA+SA facilities serving infants.

### 3.3 Platform-sensitive scale versus residual vaccine-preventable space

Table 3 and Figure 3 compare the two quantities, keeping measured values and scenario estimates strictly separated. Platform-sensitive deaths in 2023 were 222,810 for the five-pathogen opportunistic/hospital group (182,458 of them, 81.9%, in SSA+SA) and 396,995 for the full 21-pathogen platform channel (323,263, or 81.4%, in SSA+SA). Both are sums of GBD point estimates with no modeling on our side. The residual vaccine-preventable space, by contrast, is a scenario with fully disclosed arithmetic: the low scenario multiplies the 227,976.5 pneumococcal deaths of 2023 by 0.23, the smaller global uncovered fraction (Hib3), giving 52,435; the high scenario uses 0.34, the PCV3 uncovered fraction, giving 77,512; and a Hib add-on multiplies 17,774.8 Hib deaths by 0.23, giving 4,088. Against these scenarios, platform-sensitive deaths were 2.9–5.1 times as large as the residual vaccine-preventable space: 2.87 for the opportunistic/hospital group against the high scenario, 5.12 for the full channel against the high scenario, and 4.25 for the opportunistic/hospital group against the low scenario.

**Figure 3.**
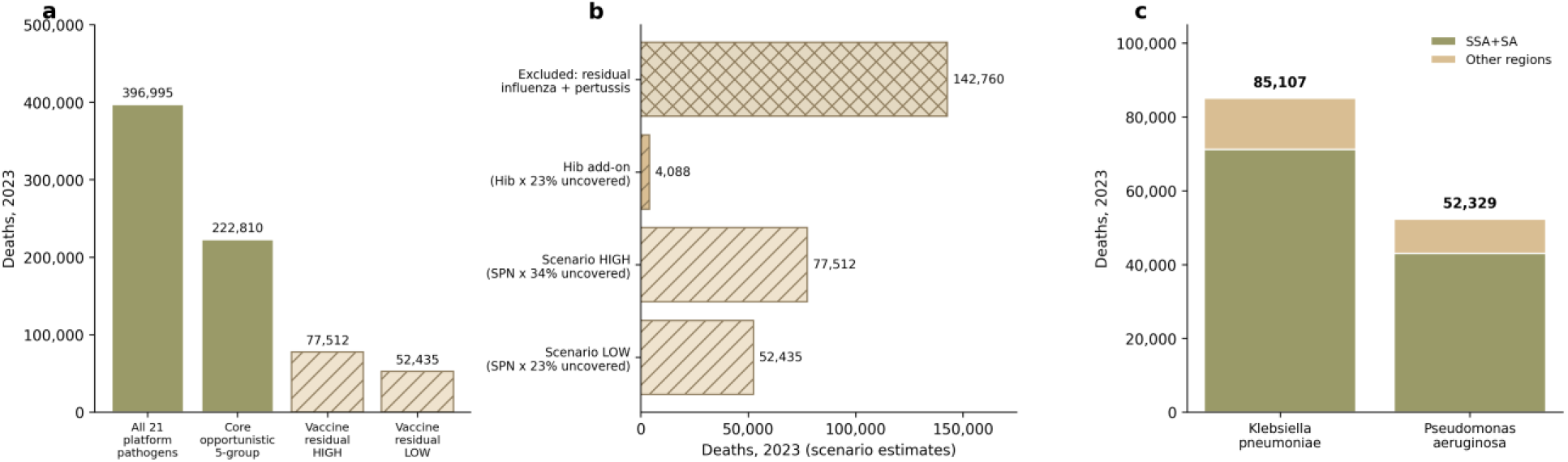
Platform scale versus vaccine residual space, 2023. (a) Measured platform-sensitive deaths (all 21 platform-channel pathogens 396,995; core opportunistic five-pathogen group 222,810) versus scenario residual vaccine-preventable space (HIGH 77,512; LOW 52,435); measured pools are 2.9–5.1 times the scenario space. (b) Scenario decomposition: LOW (SPN deaths × 23% uncovered = 52,435), HIGH (SPN deaths × 34% uncovered = 77,512), Hib add-on (Hib deaths × 23% = 4,088); residual influenza and pertussis deaths excluded from the scenario (142,760) are shown separately. Scenario assumptions: deaths distributed proportionally to coverage, full vaccine efficacy within reach; not a cost-effectiveness figure. (c) High-tier (>50,000 deaths) platform-sensitive cell: Klebsiella pneumoniae 85,107 and Pseudomonas aeruginosa 52,329 (jointly 137,436), split into SSA+SA and other regions.

**Table 3.** Platform-sensitive deaths (measured, 2023) versus residual vaccine-preventable space (scenario).

| Quantity | Deaths | SSA+SA deaths (share) | Basis |
|---|---|---|---|
| Opportunistic/hospital group (5 pathogens) | 222,810 | 182,458 (81.9%) | Measured (GBD 2023) |
| Full platform channel (21 pathogens) | 396,995 | 323,263 (81.4%) | Measured (GBD 2023) |
| Vaccine residual (pneumococcal), low scenario | 52,435 | — | 227,976.5 × 0.23 (Hib3 uncovered, conservative bracket) |
| Vaccine residual (pneumococcal), high scenario | 77,512 | — | 227,976.5 × 0.34 (PCV3 uncovered) |
| Hib add-on | 4,088 | — | 17,774.8 × 0.23 |
| Ratio: platform / residual | 2.9–5.1× | — | 2.87 (opportunistic group/high), 5.12 (full channel/high), 4.25 (opportunistic group/low) |

Scenario scope and assumptions: the scenario covers pneumococcus and Hib only; residual influenza and pertussis deaths (142,760 in 2023) are excluded. Deaths distributed in proportion to coverage; full vaccine efficacy within reach; indirect effects, serotype replacement, and higher baseline risk of uncovered populations ignored. Both the restricted scope and the baseline-risk omission bias the residual downward, so the true residual is larger and the ratios are upper bounds. Ratios order magnitudes; they are not cost-effectiveness results.

The comparison is one of magnitude, not of cost-effectiveness, and we flag the scenario scope and assumptions each time these numbers are used: the scenario covers pneumococcus and Hib only (influenza and pertussis, 142,760 deaths in 2023, are excluded); deaths are assumed to distribute in proportion to coverage; the vaccine is assumed fully effective within reach; and indirect effects, serotype replacement, and the higher baseline risk of uncovered populations are ignored. The restricted scope and the baseline-risk omission both bias the residual estimate downward, so the true vaccine residual is most plausibly larger than our figures and the 2.9–5.1 ratio is an upper bound that overstates the platform side’s relative size; the scenario is therefore not conservative with respect to the vaccine side, and we make no claim in that direction. Even with that qualification, the measured platform-sensitive pool is several-fold larger than the scenario residual and is more concentrated in poor regions (81–82% versus 78.6% for the vaccine channel). The conclusion that the marginal sequence must now engage the platform channel does not depend on the ratio being five rather than three.

### 3.4 Age tropism and poverty lock

Table 4 and Figure 4 show the 2×2 classification of the 14 computable nodes against the 80.59% all-spectrum lock reference. Nine pathogens were classified in the poverty-locked, infant-tropic cell (*Chlamydia*, group B streptococcus, *Klebsiella pneumoniae*, *Pseudomonas aeruginosa*, pneumococcus, influenza, *Escherichia coli*, *Enterobacter*, and Hib), carrying 480,922 deaths in 2023, roughly four-fifths of all deaths carried by the 14 classified nodes. Two pathogens were locked but older-tropic (*Acinetobacter baumannii* and *Mycoplasma*; 38,480 deaths); one was infant-tropic but not locked (respiratory syncytial virus; 28,052 deaths); and two were neither locked nor infant-tropic (*Staphylococcus aureus* and *Legionella*; 38,610 deaths). The cell configuration was 9/2/1/2, and the enrichment of the locked, infant-tropic cell, although large in point terms, was not statistically significant: Fisher exact odds ratio 9.0, p = 0.1758, n = 14. We report this as-is: the direction is consistent with an association between poverty lock and infant tropism, but the available nodes are too few to establish it, and we treat the 2×2 as a configuration map rather than a tested hypothesis.

**Figure 4.**
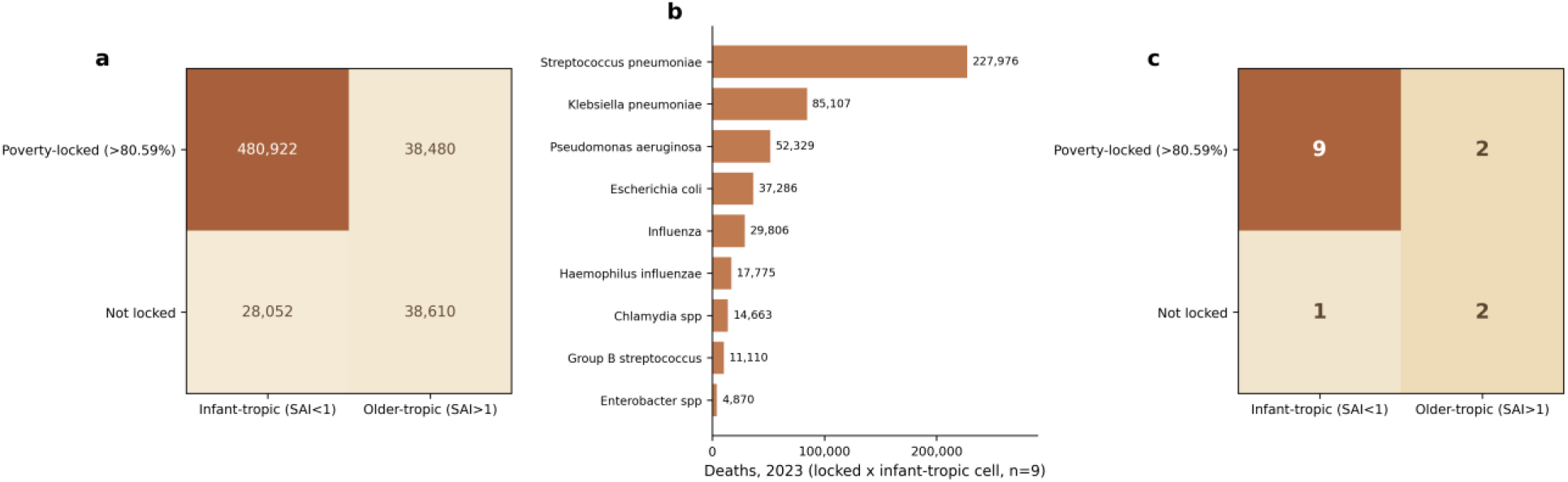
Age tropism × poverty lock for the 14 computable nodes. (a) Deaths per cell, 2023: poverty-locked × infant-tropic 480,922; locked × older-tropic 38,480; not locked × infant-tropic 28,052; not locked × older-tropic 38,610. (b) The locked × infant-tropic cell (nine pathogens, 480,922 deaths) by pathogen: Streptococcus pneumoniae 227,976; Klebsiella pneumoniae 85,107; Pseudomonas aeruginosa 52,329; Escherichia coli 37,286; influenza 29,806; Haemophilus influenzae 17,775; Chlamydia spp 14,663; group B streptococcus 11,110; Enterobacter spp 4,870. (c) Pathogen counts per cell (9/2/1/2); Fisher exact test odds ratio 9.0, p = 0.1758 (n = 14).

**Figure 5.**
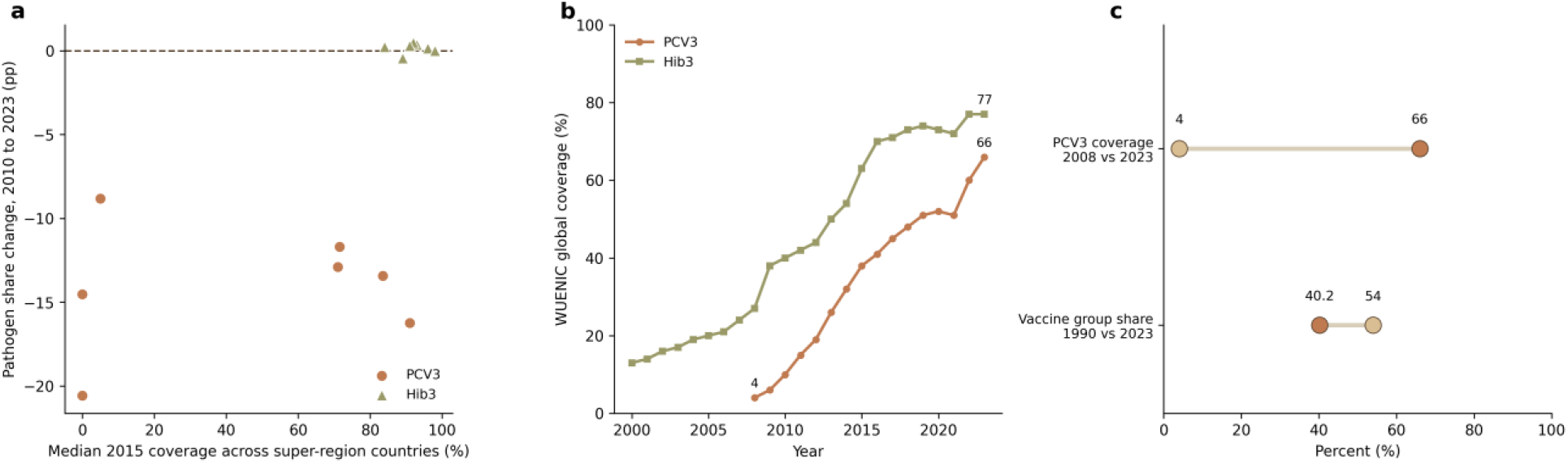
Dose–response analysis, null result reported as-is. (a) Super-region scatter (n = 7): median 2015 WUENIC coverage versus the 2010–2023 change in pathogen share; PCV3 Spearman ρ = 0.108 (p = 0.818), Hib3 ρ = −0.036 (p = 0.939) — null associations. (b) WUENIC global coverage: PCV3 4% (2008) to 66% (2023); Hib3 77% (2023). (c) Juxtaposition of the two facts: PCV3 coverage rose from 4% to 66% while the vaccine-group share fell from 54.0% to 40.2%; coverage expansion did not map linearly onto share decline across the seven super-regions.

**Table 4.** Age tropism × poverty lock, 2×2 (14 computable nodes; lock reference 80.59% SSA+SA).

|  | **Infant-tropic (SAI < 1)** | **Older-tropic (SAI ≥ 1)** |
|---|---|---|
| **Poverty-locked (>80.59%)** | 9 pathogens / 480,922 deaths (*Chlamydia*, GBS, *Klebsiella*, *Pseudomonas*, pneumococcus, influenza, *E. coli*, *Enterobacter*, Hib) | 2 pathogens / 38,480 deaths (*A. baumannii*, *Mycoplasma*) |
| **Not locked** | 1 pathogen / 28,052 deaths (RSV) | 2 pathogens / 38,610 deaths (*S. aureus*, *Legionella*) |

Fisher exact test: OR = 9.0, p = 0.1758 (direction consistent with locked × infant-tropic enrichment; not significant at n = 14, reported as-is).

### 3.5 Threshold sensitivity and bootstrap stability

The base classification is robust to downward but not upward perturbation of the lock reference (Table 5). Lowering the reference by 5 percentage points (to 75.59%) expands the locked set from 19 to 27 of 29 nodes and the locked × infant-tropic cell from 9 to 10 of the 14 classifiable nodes (cell deaths 480,922 to 508,974), leaving the qualitative reading unchanged. Raising it by 5 points (to 85.59%) empties the locked cell: only tuberculosis and *Chlamydia* spp. exceed 85.59% among all 29 nodes, and among the 14 classifiable nodes only *Chlamydia* spp. remains locked, because the locked cluster sits immediately above the reference (11 of the 14 classifiable nodes lie between 80.6% and 86.0% SSA+SA share). Perturbing the SAI threshold by ±5% relative (0.95 or 1.05) leaves the 2×2 unchanged at the upper bound—no node has an SAI in (1, 1.05]— and shifts it to 6/5/1/2 at the lower bound (Fisher OR 2.4, p = 1.0). Across joint perturbations the locked × infant-tropic cell spans 0 to 10 nodes (0 to 508,974 deaths) ; no scenario reached p < 0.05 (p 0.176–1.0 across the nine scenarios), and in the four scenarios where the odds ratio is undefined (an empty cell) p is 0.286 or 1.0. In 500 bootstrap resamples of the 14 nodes (seed 42; Additional file 7), the Fisher odds ratio varied widely: median 5.0 (P2.5–P97.5 0.0–27.0) among the 281 of 500 replicates with a defined odds ratio; p < 0.05 in 19.0% of all 500 replicates (5.7% among the defined-OR replicates); and the odds ratio undefined in 219 replicates (43.8%) through an empty locked × older-tropic or not-locked × infant-tropic cell. Because each node’s locked/infant status is a deterministic function of its two indices, resampling nodes cannot alter any label; set-membership stability is therefore not an estimable quantity here, and the bootstrap is reported only for the Fisher enrichment statistic. The enrichment estimate, not the classification, is the fragile element—consistent with the underpowered-test caveat above.

**Table 5.** Threshold sensitivity of the age-tropism × poverty-lock 2×2 classification (14 computable nodes) and of the 29-node locked set.

| Scenario | Locked nodes (of 29) | Locked × infant-tropic | Locked × older-tropic | Not locked × infant-tropic | Not locked × older-tropic | Fisher OR | p value |
|---|---|---|---|---|---|---|---|
| **Base (lock 80.59%; SAI 1.00)** | 19 | 9 / 480,922 | 2 / 38,480 | 1 / 28,052 | 2 / 38,610 | 9.0 | 0.1758 |
| **Lock 75.59% (−5 pp)** | 27 | 10 / 508,974 | 3 / 76,497 | 0 / 0 | 1 / 594 | — | 0.2857 |
| **Lock 85.59% (+5 pp)** | 2 | 1 / 14,663 | 0 / 0 | 9 / 494,311 | 4 / 77,091 | — | 1.0000 |
| **SAI 0.95 (−5% relative)** | 19 | 6 / 450,279 | 5 / 69,123 | 1 / 28,052 | 2 / 38,610 | 2.4 | 1.0000 |
| **SAI 1.05 (+5% relative)** | 19 | 9 / 480,922 | 2 / 38,480 | 1 / 28,052 | 2 / 38,610 | 9.0 | 0.1758 |
| **Joint: lock 75.59% × SAI 0.95** | 27 | 7 / 478,331 | 6 / 107,140 | 0 / 0 | 1 / 594 | — | 1.0000 |
| **Joint: lock 75.59% × SAI 1.05** | 27 | 10 / 508,974 | 3 / 76,497 | 0 / 0 | 1 / 594 | — | 0.2857 |
| **Joint: lock 85.59% × SAI 0.95** | 2 | 0 / 0 | 1 / 14,663 | 7 / 478,331 | 6 / 93,070 | 0.0 | 1.0000 |
| **Joint: lock 85.59% × SAI 1.05** | 2 | 1 / 14,663 | 0 / 0 | 9 / 494,311 | 4 / 77,091 | — | 1.0000 |

The configuration is nonetheless decision-relevant. The locked, infant-tropic cell contains three vaccine-reachable pathogens (pneumococcus, influenza, and Hib), one mixed pathogen (group B streptococcus), and five platform-sensitive pathogens; it is the one cell that both intervention channels must jointly cover. For its vaccine-free members (*Klebsiella pneumoniae*, *Pseudomonas aeruginosa*, *Escherichia coli*, *Enterobacter*, and *Chlamydia*), platform interventions are not the second-best but the only currently available channel. Conversely, the pathogens outside this cell are either geographically dispersed (RSV, *Staphylococcus aureus*, *Legionella*) or older-tropic (*Acinetobacter baumannii*, *Mycoplasma*), and none carries a death count approaching the locked cell’s. The practical output of the matrix is therefore a single, tightly specified delivery target: infants in sub-Saharan Africa and South Asia, reached through a combined vaccine-completion and platform-capability package.

Cell entries are node count / deaths in 2023. Lock reference perturbed ±5 percentage points from 80.59%; SAI threshold perturbed ±5% relative from 1.00 (SAI is a ratio, so the relative perturbation is the applicable analogue of the percentage-point rule). ‘—’ = Fisher odds ratio undefined (empty cell). Share tiers (>50,000; 5,000–50,000; <5,000 deaths) are absolute-count cutoffs and the intervention-channel assignment is a non-numeric rule, so neither enters the percentage-point design. Bootstrap (500 resamples of the 14 nodes, seed 42; Additional file 7): because each node’s locked/infant status is a deterministic function of its two indices, resampling cannot alter any label, so set-membership stability is not an estimable quantity and the bootstrap is reported only for the Fisher enrichment statistic — OR median 5.0 (P2.5–P97.5 0.0–27.0) among the 281 of 500 replicates with a defined OR; p < 0.05 in 19.0% of all 500 replicates (5.7% among defined-OR replicates); OR undefined in 219 replicates (43.8%) through an empty locked × older-tropic or not-locked × infant-tropic cell.

## 4. Discussion

### 4.1 The second half of the vaccine era

The first half of the vaccine era in child pneumonia was defined by closing introduction and coverage gaps for Hib and PCV. That agenda is unfinished but advanced: global PCV3 coverage reached 66% and Hib3 coverage 77% by 2023 ^4^, and the vaccine group share has fallen to approximately two-fifths of the spectrum (40.2%) under the primary caliber. The second half will be defined by managing a diversified residual spectrum in which 41.1% of deaths now arise from pathogens with no licensed childhood vaccine. Our results quantify the structural case for rebalancing the marginal resource: platform-sensitive deaths outnumber the scenario estimate of residual vaccine-preventable deaths by a factor of roughly three to five (an upper-bound ratio, given the scenario’s restricted pathogen scope), are more concentrated in sub-Saharan Africa and South Asia (81.4% versus 78.6%), and cluster in infants—the age band in which severe disease, hypoxaemia, and case fatality peak. None of this diminishes the vaccine achievement; it describes its consequence. The better the vaccine era performs, the larger the platform-sensitive share of what remains.

The platform intervention list is short and evidence-anchored (Table 6). First, oxygen systems. Hypoxaemia is a common complication of childhood pneumonia and acute lower respiratory infection and a recognized risk factor for death, yet it was long overlooked in pneumonia control strategies, which historically emphasized case finding and antibiotics over supportive care ^6,7^. The interventional evidence, although not randomized, is unusually consistent. In a multihospital effectiveness study across five hospitals in Papua New Guinea, improved oxygen systems (concentrators, pulse oximetry, and clinical protocols) reduced pneumonia case fatality from 4.97% to 3.22%, a 35% lower risk of death (RR 0.65, 95% CI 0.52–0.78), at a reported cost of US$51 per patient treated, US$1,673 per life saved, and US$50 per DALY averted ^11^. A systematic review and meta-analysis spanning 75 hospitals found a pooled odds ratio of 0.52 (95% CI 0.39– 0.70) for childhood pneumonia mortality after oxygen systems strengthening, at a median of US$62 per DALY averted (range 44–225); the authors rate the certainty of evidence as low because the underlying designs were non-randomized, and we carry that rating forward without embellishment ^12^. WHO’s manual for health workers codifies hypoxaemia detection by pulse oximetry and oxygen delivery at facility level, giving the intervention a normative anchor ^13^. The Lancet Global Health Commission on Medical Oxygen Security has since quantified the investment case at scale: closing the acute medical and surgical oxygen coverage gap in low- and middle-income countries requires an additional US$6.8 billion per year (US$34 billion over 2025–2030), and the Commission judges medical oxygen to be as cost-effective as routine childhood immunization^16^. Second, antimicrobial access and stewardship. The platform-sensitive pathogens include leading contributors to the global burden of bacterial antimicrobial resistance (AMR) ^17^; access programs that are not paired with stewardship will erode their own effectiveness, so the two must be financed as one line item. Third, infection prevention and control, which targets the hospital-associated component of the spectrum (*Klebsiella*, *Pseudomonas*, *Acinetobacter*, *Staphylococcus*) at its point of transmission, particularly in neonatal units. Fourth, severe-case referral and grading, which determines whether hypoxaemic children reach oxygen-capable facilities in time for the oxygen to matter. Fifth, nutrition, a cross-cutting determinant of pneumonia incidence, severity, and case fatality that no pathogen-specific tool touches. The unifying property of the list is that none of these interventions is pathogen- specific: all act on the shared severe-disease pathway, which is precisely why their relative value rises as the spectrum diversifies and why their returns do not depend on which pathogen happens to dominate next.

**Table 6.** Literature anchors for platform interventions (published values, original certainty ratings retained).

| Intervention | Evidence | Key published figures | Certainty |
|---|---|---|---|
| Oxygen systems (concentrators, pulse oximetry, protocols) | Duke et al., Lancet 2008, multihospital effectiveness study, Papua New Guinea<sup>12</sup> | Pneumonia CFR 4.97% to 3.22%; RR 0.65 (95% CI 0.52–0.78); US\$51 per patient treated; US\$1,673 per life saved; US\$50 per DALY averted | Non-randomized effectiveness study |
| Oxygen systems strengthening | Lam et al., BMJ Glob Health 2021, systematic review/meta-analysis, 75 hospitals<sup>13</sup> | Pooled OR 0.52 (95% CI 0.39–0.70) for childhood pneumonia mortality; median US\$62 per DALY averted (range 44–225) | Low (non-randomized designs), as rated by authors |
| Hypoxaemia burden and detection | Subhi et al., Lancet Infect Dis 2009<sup>6</sup>; Rahman et al., Lancet Glob Health 2022<sup>7</sup>; WHO manual 2016<sup>14</sup> | Hypoxaemia common in childhood ALRI and a recognized risk factor for death, long overlooked in control strategies; pulse oximetry-based detection and oxygen delivery codified for health workers | Systematic reviews; normative guidance |
| Antimicrobial access and stewardship | Antimicrobial Resistance Collaborators, Lancet 2022<sup>18</sup> | Platform-sensitive pathogens are leading contributors to the global AMR burden, requiring access paired with stewardship | Systematic analysis |

### 4.2 Sequencing, not substitution

The argument is for a relay, not a replacement. The scenario residual vaccine-preventable space of 52,435–77,512 deaths (plus a Hib add-on of 4,088) remains large, certain, and inexpensive relative to most alternatives; closing coverage gaps is the highest-certainty component of any sequence, and nothing in our results argues for diverting funds from vaccine completion. Three features of the marginal structure, however, argue that what comes *next* after each additional vaccine dollar is increasingly a platform dollar. First, the two channels are not in competition for the same deaths: the residual vaccine-preventable pool will shrink with coverage regardless of what platforms do, while the platform-sensitive pool is untouched by immunization. The comparison in Table 3 therefore reads as “additional addressable pool,” not “either/or.” Second, vaccine returns decline at the margin as coverage saturates and as the vaccine-preventable share of the spectrum falls (54.0% to 40.2% since 1990), whereas platform returns do not depend on pathogen identity and rise mechanically as the share of the spectrum that only platforms can reach grows; that share already stands at 41.1% for the full platform channel in 2023, and the five-pathogen opportunistic/hospital group within it rose from 18.1% to 23.1%. Third, the dose– response null we report reinforces the sequencing point from the empirical side: because regional coverage levels do not explain regional share declines, the residual burden after vaccination appears to be mediated by structural factors (treatment access, facility capability, nutrition) that only platform investment reaches. A prudent sequence therefore front-loads vaccine completion, shifts the marginal dollar progressively toward platform capability in the regions where the residual burden is locked, and sustains vaccine programs throughout as the floor of the system. The serotype-replacement literature points in the same direction: pneumococcal conjugate vaccines clear vaccine serotypes while non-vaccine serotypes expand into the vacated ecological space ^14,18^, so part of the pneumococcal residual is structurally platform-sensitive even within the vaccine channel’s flagship pathogen. Formal economic evaluation of the sequence (comparing marginal cost per death averted across the two channels under local prices) is a necessary next step that this framework is designed to inform rather than replace.

### 4.3 Poverty lock, tropism, and package design

The configuration matrix gives the sequence a geographic and demographic address. Nine of 14 classifiable pathogens (including the high-burden vaccine-reachable pathogens and both high-share platform-sensitive pathogens) are simultaneously infant-tropic and locked to SSA+SA, carrying 480,922 deaths in 2023. Because the lock reference (80.59%) is the deaths-weighted average for the whole spectrum, the finding is that the largest pathogens are concentrated *above* an already high average: the residual burden is not merely poor-region but is specifically the burden of poor-region infancy. For this cell, the efficient product is a package rather than a project: routine immunization completion, an oxygen-and-pulse-oximetry capability with referral grading, antimicrobial access with stewardship, infection prevention and control in neonatal units, and nutrition support, designed around the newborn and infant contact schedule (delivery, postnatal, and immunization visits) that SSA+SA health systems already operate. The contact schedule matters practically: the same facility visits that deliver the remaining vaccine doses are the natural delivery points for pulse-oximetry screening, danger-sign triage, and referral, so the incremental cost of the platform components is partly a cost of capability, not of an entirely new delivery channel. The near-uniformity of poverty concentration across matrix cells (Table 2) means this package design does not need to be re-targeted by pathogen; one geography, one age band, two channels.

### 4.4 Limitations

Four limitations qualify these findings. First, channel classification is a rule-based judgment; the mixed assignment of tuberculosis and group B streptococcus could reasonably be argued otherwise, and channel totals would shift modestly under alternative assignments. Second, the vaccine-residual figures are scenarios under stated assumptions, not measurements; the scenario covers pneumococcus and Hib only, omitting residual influenza and pertussis deaths (142,760 deaths in 2023) together with the higher baseline risk of uncovered populations, so the true residual is larger and the 2.9–5.1 platform-to-residual ratio is an upper bound rather than a conservative estimate of the platform side’s relative scale. The platform-versus-vaccine comparison is an ordering of magnitudes: the framework is a prioritization tool, not a cost-effectiveness calculation. Specifically, the 2.9–5.1× pool-size ratio compares the sizes of two addressable death pools, not their marginal cost-effectiveness—pool scale alone does not establish where the next dollar buys the most health—and the ordering it supports is a prioritization framework for structuring the pricing exercise, not a resource-allocation recommendation. Third, GBD estimates are modeled quantities with uncertainty intervals that we do not propagate through the matrix; point-estimate rankings should be read accordingly. Fourth, the Fisher test of tropism–lock enrichment is underpowered at n = 14 (OR = 9.0 but p = 0.1758), and the dose–response analysis is an ecological super-region comparison at n = 7 that cannot exclude country-level coverage effects; we present both as directional rather than confirmatory. Two further caveats deserve emphasis. The literature anchors for oxygen systems rest on non-randomized designs with low certainty ratings ^11,12^, and we have not converted their effect sizes into deaths-avertible projections for our matrix cells; doing so would be a legitimate modeling exercise but a different paper. Finally, the framework is calibrated to the global and super-region scale at which our inputs are measured; country-level sequencing decisions require country-level burden, coverage, and price data, and the matrix should be read as a template for that exercise rather than a verdict on any specific national allocation. The threshold sensitivity analysis quantifies the fragility of the tropism–lock reading directly: it survives a 5-percentage-point lowering of the lock reference but not a 5-point increase, because the locked cluster lies immediately above the reference line (11 of 14 classifiable nodes between 80.6% and 86.0%), and the bootstrap confirms that the Fisher enrichment, not the classification itself, is the unstable element (Table 5).

## 5. Conclusions

Child pneumonia control is entering the second half of the vaccine era. Between 1990 and 2023 the pathogen spectrum diversified from the equivalent of fewer than six effective pathogens to nearly ten (effN 5.57 to 9.94), the vaccine-preventable group share fell from 54.0% to 40.2%, and the residual burden shifted toward platform-sensitive, poverty-locked, infant-tropic pathogens. In 2023, the 21 platform-sensitive pathogens carried 396,995 deaths in ages 0–19 years (2.9–5.1 times an upper-bound scenario estimate of residual pneumococcal-and-Hib deaths) and 81.4% of those deaths occurred in sub-Saharan Africa and South Asia. The empirical record also carries a warning against over-reading coverage alone: super-region vaccine coverage showed no significant association with pathogen-share decline (PCV3 ρ = 0.108, p = 0.818; Hib3 ρ = −0.036, p = 0.939), indicating that the residual burden is mediated by structural factors that immunization does not reach.

The policy implication is sequencing, not substitution. Vaccine completion remains the highest-certainty investment on the table (52,435–77,512 residual deaths under stated scenario assumptions) and should be finished, not abandoned. But the next marginal dollar should increasingly fund platform capabilities: oxygen systems with pulse oximetry, antimicrobial access with stewardship, infection prevention and control, severe-case referral, and nutrition, delivered as an integrated package to the infant populations of the locked regions through the contact points that already exist. This framework prioritizes; it does not price. The next step is formal economic evaluation of the sequence, with channel classifications and scenario assumptions subjected to the sensitivity analysis that local decision-making requires.

## Data Availability

Availability of data and materials: All input estimates are publicly available from the GBD 2023 Results Tool (https://vizhub.healthdata.org/gbd-results/) and from WHO/UNICEF Estimates of National Immunization Coverage via the WHO Global Health Observatory (https://data.who.int). The analysis tables reproducing every figure and table in this study are available from the corresponding author on reasonable request. The country-level dataset of pathogen-attributable deaths by intervention channel (204 countries, five time points 1990-2023) is provided as Additional file 2 (03_Dataset.xlsx). The threshold-sensitivity grid and bootstrap-stability output added in this revision are provided as Additional file 7 (sensitivity_pm5pp_PneumoniaPlatform.csv and bootstrap_stability_PneumoniaPlatform.csv) with the fully reproducible analysis script (code/threshold_sensitivity_bootstrap.py).

## Declarations

### Ethics approval and consent to participate

Not applicable. This study is a secondary analysis of publicly available, de-identified modelled estimates (Global Burden of Disease Study 2023) and published immunization-coverage aggregates (WUENIC); no individual-level data were used.

### Consent for publication

Not applicable.

### Availability of data and materials

All input estimates are publicly available from the GBD 2023 Results Tool (https://vizhub.healthdata.org/gbd-results/) and from WHO/UNICEF Estimates of National Immunization Coverage via the WHO Global Health Observatory (https://data.who.int). The analysis tables reproducing every figure and table in this study are available from the corresponding author on reasonable request. The country-level dataset of pathogen-attributable deaths by intervention channel (204 countries, five time points 1990–2023) is provided as Additional file 2 (03_Dataset.xlsx). The threshold-sensitivity grid and bootstrap-stability output added in this revision are provided as Additional file 7 (sensitivity_pm5pp_PneumoniaPlatform.csv and bootstrap_stability_PneumoniaPlatform.csv) with the fully reproducible analysis script (code/threshold_sensitivity_bootstrap.py).

### Funding

This work was supported by the Beijing Science and Technology Nova Program Interdisciplinary Project (20230484439) and the Beijing High-Level Public Health Technical Talents Project (2023-02-33). The funder had no role in study design, data collection, data analysis, data interpretation, or writing of the report.

Presentation This work has not been presented at any scientific meeting.

Disclosure The authors declare no conflicts of interest. AI tools were used for language editing and for assistance with code generation and modification; the authors take full responsibility for the content.

### Authors’ contributions

SC conceptualised the study, supervised the analysis and drafted the manuscript; DL, HC and XW curated the GBD 2023 extracts, recomputed the spectrum tabulations and prepared the figures and tables; HC and DL performed the coverage dose– response and tropism–lock analyses and verified the literature anchors against PubMed records; CS contributed to interpretation and critical revision of the manuscript. All authors read and approved the final manuscript.

## Acknowledgements

The authors thank the Institute for Health Metrics and Evaluation and the Global Burden of Disease collaborative network, and the WHO/UNICEF WUENIC team, for making the underlying estimates publicly available.

## Additional files

Additional file 1: Supplementary_Materials.docx — Supplementary Tables S1–S8 (per-pathogen channel classification matrix; tropism–lock exclusion list; SAI/LAI definitions; dual-caliber transition table; scenario arithmetic; effN and tier shares; dose–response results; country-level channel composition, 2023). Additional files 3–6: Figures 1–5 as separate PNG and TIFF files (300 dpi).

Additional file 2: 03_Dataset.xlsx — country-level dataset of pathogen-attributable deaths by intervention channel (204 countries, 26 aetiologies, five time points 1990–2023), including channel classification, population denominators and crude death rates per 100,000 population aged 0–19 years.

Additional file 3: Threshold sensitivity

