## Supplementary_Materials for "After the vaccine era: sequencing platform investments as the childhood pneumonia spectrum diversifies"

Contents: Supplementary Tables S1–S8 (this file). Additional file 2: 03_Dataset.xlsx — country- level dataset of pathogen-attributable deaths by intervention channel (204 countries, five time points 1990–2023).

### Supplementary Table S1. Per-pathogen intervention-channel classification matrix: all 29 pathogens, dominant channel, classification rationale, deaths in 2023 (ages 0–19, global), and sub-Saharan Africa plus South Asia (SSA+SA) share of deaths

| Pathogen | Type | Dominant channel | Classification rationale | Deaths 2023 | SSA+SA share (%) |
|---|---|---|---|---|---|
| Streptococcus pneumoniae | Bacterium | Vaccine- reachable | Licensed PCV in EPI; residual deaths track coverage gap (WUENIC PCV3) | 227,976 | 81.7 |
| Whooping cough | Bacterium | Vaccine- reachable | Licensed pertussis-containing vaccine (DTP) in EPI | 112,954 | 82.7 |
| Tuberculosis | Bacterium | Mixed (vaccine + platform) | BCG gives partial protection against severe childhood TB; control mainly via case-finding/treat ment platform | 87,764 | 87.1 |
| Klebsiella pneumoniae | Bacterium | Platform- sensitive | No licensed pediatric vaccine; prevention/treatme nt via platform (oxygen, antimicrobial access, infection prevention & control, critical-care referral) | 85,107 | 83.6 |
| COVID-19 | Virus | Vaccine- reachable | Licensed SARS-CoV-2 vaccines; pediatric priority low | 52,899 | 54.0 |
| Pseudomonas aeruginosa | Bacterium | Platform- sensitive | No licensed pediatric vaccine; prevention/treatme nt via platform (oxygen, antimicrobial access, infection prevention & control, critical-care referral) | 52,329 | 82.2 |

| Pathogen | Type | Dominant channel | Classification rationale | Deaths 2023 | SSA+SA share (%) |
|---|---|---|---|---|---|
| Staphylococcus aureus | Bacterium | Platform- sensitive | No licensed pediatric vaccine; prevention/treatment via platform (oxygen, antimicrobial access, infection prevention & control, critical-care referral) | 38,016 | 76.6 |
| Escherichia coli | Bacterium | Platform- sensitive | No licensed pediatric vaccine; prevention/treatment via platform (oxygen, antimicrobial access, infection prevention & control, critical-care referral) | 37,286 | 80.9 |
| Other Mycobacterium species (non-TB, non-Leprosy) | Bacterium | Platform- sensitive | No licensed pediatric vaccine; prevention/treatment via platform (oxygen, antimicrobial access, infection prevention & control, critical-care referral) | 35,259 | 82.3 |
| Influenza | Virus | Vaccine- reachable | Licensed seasonal influenza vaccines | 29,806 | 81.1 |
| Respiratory syncytial virus | Virus | Mixed (vaccine + platform) | Maternal RSV vaccine & long-acting mAb newly licensed; care remains supportive (oxygen platform) | 28,052 | 79.3 |
| Mycoplasma | Bacterium | Platform- sensitive | No licensed pediatric vaccine; prevention/treatment via platform (oxygen, antimicrobial access, infection prevention & control, critical-care referral) | 26,382 | 81.6 |

| Pathogen | Type | Dominant channel | Classification rationale | Deaths 2023 | SSA+SA share (%) |
|---|---|---|---|---|---|
| Group A Streptococcus | Bacterium | Platform- sensitive | No licensed pediatric vaccine; prevention/treatment via platform (oxygen, antimicrobial access, infection prevention & control, critical-care referral) | 19,333 | 77.0 |
| Other gram-negative bacteria | Bacterium | Platform- sensitive | No licensed pediatric vaccine; prevention/treatment via platform (oxygen, antimicrobial access, infection prevention & control, critical-care referral) | 19,079 | 82.9 |
| Haemophilus influenzae | Bacterium | Vaccine- reachable | Licensed Hib vaccine in EPI (WUENIC Hib3) | 17,775 | 80.6 |
| Other bacterial and viral pathogens | Other (mixed bacterial/viral) | Platform- sensitive | No licensed pediatric vaccine; prevention/treatment via platform (oxygen, antimicrobial access, infection prevention & control, critical-care referral) | 16,173 | 83.0 |
| Chlamydia spp | Bacterium | Platform- sensitive | No licensed pediatric vaccine; prevention/treatment via platform (oxygen, antimicrobial access, infection prevention & control, critical-care referral) | 14,663 | 86.0 |
| Acinetobacter baumannii | Bacterium | Platform- sensitive | No licensed pediatric vaccine; prevention/treatment via platform (oxygen, antimicrobial access, infection prevention & control, critical-care referral) | 12,098 | 84.3 |

| Pathogen | Type | Dominant channel | Classification rationale | Deaths 2023 | SSA+SA share (%) |
|---|---|---|---|---|---|
| Aspergillus spp. | Fungus | Platform- sensitive | No licensed pediatric vaccine; prevention/treatment via platform (oxygen, antimicrobial access, infection prevention & control, critical-care referral) | 11,781 | 76.8 |
| Group B streptococcus | Bacterium | Mixed (vaccine + platform) | Maternal GBS vaccines in late-stage trials; intrapartum antibiotic prophylaxis is the current platform | 11,110 | 83.8 |
| Other fungi | Fungus | Platform- sensitive | No licensed pediatric vaccine; prevention/treatment via platform (oxygen, antimicrobial access, infection prevention & control, critical-care referral) | 7,496 | 79.3 |
| Other Klebsiella species | Bacterium | Platform- sensitive | No licensed pediatric vaccine; prevention/treatment via platform (oxygen, antimicrobial access, infection prevention & control, critical-care referral) | 5,690 | 79.3 |
| Enterobacter spp | Bacterium | Platform- sensitive | No licensed pediatric vaccine; prevention/treatment via platform (oxygen, antimicrobial access, infection prevention & control, critical-care referral) | 4,870 | 80.9 |

| Pathogen | Type | Dominant channel | Classification rationale | Deaths 2023 | SSA+SA share (%) |
|---|---|---|---|---|---|
| Other Acinetobacter species | Bacterium | Platform- sensitive | No licensed pediatric vaccine; prevention/treatment via platform (oxygen, antimicrobial access, infection prevention & control, critical-care referral) | 4,143 | 82.6 |
| Serratia spp. | Bacterium | Platform- sensitive | No licensed pediatric vaccine; prevention/treatment via platform (oxygen, antimicrobial access, infection prevention & control, critical-care referral) | 2,782 | 80.7 |
| Citrobacter spp. | Bacterium | Platform- sensitive | No licensed pediatric vaccine; prevention/treatment via platform (oxygen, antimicrobial access, infection prevention & control, critical-care referral) | 1,666 | 76.1 |
| Proteus spp. | Bacterium | Platform- sensitive | No licensed pediatric vaccine; prevention/treatment via platform (oxygen, antimicrobial access, infection prevention & control, critical-care referral) | 1,304 | 78.4 |
| Morganella spp. | Bacterium | Platform- sensitive | No licensed pediatric vaccine; prevention/treatment via platform (oxygen, antimicrobial access, infection prevention & control, critical-care referral) | 942 | 83.0 |

| Pathogen | Type | Dominant channel | Classification rationale | Deaths 2023 | SSA+SA share (%) |
|---|---|---|---|---|---|
| Legionella spp | Bacterium | Platform- sensitive | No licensed pediatric vaccine; prevention/treatment via platform (oxygen, antimicrobial access, infection prevention & control, critical-care referral) | 594 | 29.8 |

Note: Channel assignment follows the explicit rules in Methods §2.3: vaccine-reachable = licensed vaccine in routine immunization programs (n = 5); mixed = partial or emerging immunization with a dominant delivery-platform component (n = 3); platform-sensitive = no licensed childhood vaccine (n = 21). Deaths are GBD 2023 point estimates, ages 0–19 years, global; SSA+SA share is the combined sub-Saharan Africa and South Asia super-region share of each pathogen's 2023 deaths (all-spectrum reference 80.59%). The matrix is a prioritization tool, not a cost-effectiveness calculation. Assignment is a rule-based judgment of the dominant current channel, not a claim about future licensability.

### Supplementary Table S2. The 15 spectrum nodes excluded from the age-tropism × poverty-lock analysis (14 of 29 nodes classifiable), with exclusion reasons

| Excluded node | Exclusion category | Reason |
|---|---|---|
| Tuberculosis | Cause-level addition to the 29-node spectrum | Not part of the GBD 2021 lower respiratory infection (LRI) etiology incidence extract from which the susceptibility-age index (SAI) is computed; no pathogen- level LRI incidence age split available. |
| COVID-19 | Cause-level addition to the 29-node spectrum | As above; no pathogen-level LRI incidence age split (node absent before 2020). |
| Whooping cough | Cause-level addition to the 29-node spectrum | As above; no pathogen-level LRI incidence age split available. |
| Other Mycobacterium species (non-TB, non- Leprosy) | Aggregate/residual node | Heterogeneous residual grouping without a single- pathogen age structure; tropism indices not interpretable. |
| Other bacterial and viral pathogens | Aggregate/residual node | As above. |
| Other fungi | Aggregate/residual node | As above. |
| Other gram-negative bacteria | Aggregate/residual node | As above. |
| Other Klebsiella species | Aggregate/residual node | As above. |
| Other Acinetobacter species | Aggregate/residual node | As above. |
| Aspergillus spp. | Single pathogen absent from the index source extract | GBD 2023 rei-level addition not present in the GBD 2021 LRI etiology incidence extract; SAI not computable. |
| Group A Streptococcus | Single pathogen absent from the index source extract | As above. |

| Excluded node | Exclusion category | Reason |
|---|---|---|
| Serratia spp. | Single pathogen absent from the index source extract | As above. |
| Citrobacter spp. | Single pathogen absent from the index source extract | As above. |
| Proteus spp. | Single pathogen absent from the index source extract | As above. |
| Morganella spp. | Single pathogen absent from the index source extract | As above. |

Note: A node entered the tropism × lock analysis only if both age-tropism indices (SAI and LAI; Supplementary Table S3) were computable. The 15 excluded nodes were generated by cross- checking the 29-node spectrum against the tropism-index source table (pathogen_age_tropism_SAI_LAI_n14.csv, 14 evaluable pathogens): nodes outside the LRI etiology framework at cause level (3), aggregate/residual groupings (6), and single pathogens absent from the incidence extract underlying the SAI (6). All 15 nodes retain channel, deaths, and SSA+SA share in Supplementary Table S1 and in the main Tables 2–3; only the tropism × lock classification (main Table 4) is restricted to the 14 classifiable nodes.

### Supplementary Table S3. Age-tropism indices (SAI and LAI): definitions, computation,

provenance, and values for the 14 evaluable pathogens

Definitions. The susceptibility-age index (SAI) and lethality-age index (LAI) are share-based age-tropism indices ported from the spectrum29/spectrum26 analysis package (source table: pathogen_age_tropism_SAI_LAI_n14.csv; definitions as declared in the spectrum26 Methods, §perturbation–tropism analysis). SAI = incident cases at ages 5–14 years ÷ incident cases under 5 years, computed from GBD 2021 LRI etiology pathogen-level incidence (year 2021, global, both sexes); SAI < 1 indicates concentration of susceptibility in the youngest ages (infant-tropic). LAI = mean of the death shares at ages 15–19 and 20–24 years ÷ death share under 5 years, computed from GBD 2023 pathogen-level deaths by single age group (2023, global); LAI > 1 indicates a shift of lethality toward older ages. The quadrant with SAI ≥ 1 and LAI ≥ 1 is termed "older-concordant". In the present study the tropism axis uses SAI only (SAI < 1 = infant-tropic; SAI > 1 = older-tropic), and the poverty-lock axis uses the 29-node SSA+SA reference of 80.59%.

| Pathogen | SAI | LAI | Tropism classification (this study) |
|---|---|---|---|
| Respiratory syncytial virus | 0.0856 | 0.1713 | Infant-tropic (SAI<1) |
| Haemophilus influenzae | 0.5381 | 0.6167 | Infant-tropic (SAI<1) |
| Escherichia coli | 0.6409 | 0.7316 | Infant-tropic (SAI<1) |
| Klebsiella pneumoniae | 0.7018 | 0.7670 | Infant-tropic (SAI<1) |
| Pseudomonas aeruginosa | 0.8359 | 0.8244 | Infant-tropic (SAI<1) |
| Influenza | 0.9098 | 1.2081 | Infant-tropic (SAI<1) |
| Streptococcus pneumoniae | 0.9172 | 0.9276 | Infant-tropic (SAI<1) |
| Enterobacter spp | 0.9651 | 0.8093 | Infant-tropic (SAI<1) |
| Chlamydia spp | 0.9929 | 0.8748 | Infant-tropic (SAI<1) |
| Group B streptococcus | 0.9977 | 0.9057 | Infant-tropic (SAI<1) |
| Staphylococcus aureus | 1.2025 | 1.3728 | Older-tropic (SAI>1) |
| Mycoplasma | 1.3496 | 1.4378 | Older-tropic (SAI>1) |
| Acinetobacter baumannii | 3.4388 | 1.6922 | Older-tropic (SAI>1) |
| Legionella spp | 5.0873 | 6.7197 | Older-tropic (SAI>1) |

Note: n = 14 pathogens were evaluable; the 15 excluded nodes and reasons are listed in

Supplementary Table S2. Index values are ported unchanged from the spectrum26/spectrum29 analysis packages; the poverty-lock classification in this table was recomputed at the 29-node 80.59% reference (main Methods §2.5).

#### Supplementary Table S4. Spectrum transition under the two pathogen calibers, four time points (ages 0–19, global): full tabulation

| Year | Total deaths (29- node) | Vaccine group share, 29- caliber (%) | Opportu nistic/ hospital share, 29- caliber (%) | Other pathoge ns share, 29- caliber (%) | Vaccine group + COVID- 19 share (%) | Vaccine trio share, 26- caliber (%) | Opportu nistic share, 26- caliber (%) | effN (29- node) |
|---|---|---|---|---|---|---|---|---|
| 1990 | 2,573,48 9 | 54.0 | 18.1 | 27.9 | — | 54.6 | 22.8 | 5.57 |
| 2010 | 1,324,68 5 | 51.9 | 18.8 | 29.3 | — | 52.0 | 24.0 | 6.13 |
| 2019 | 1,042,33 6 | 44.9 | 22.1 | 33.1 | — | 41.9 | 28.2 | 8.94 |
| 2023 | 965,330 | 40.2 | 23.1 | 36.7 | 45.7 | 38.7 | 31.3 | 9.94 |

Note: Vaccine group (29-caliber) = Streptococcus pneumoniae + Haemophilus influenzae + influenza + pertussis; the +COVID-19 column adds COVID-19 as an alternative 2023 caliber. Opportunistic/hospital group = Pseudomonas aeruginosa, Staphylococcus aureus, non- tuberculous mycobacteria, Acinetobacter baumannii, Klebsiella pneumoniae. Vaccine trio (26- caliber) = pneumococcus + Hib + influenza against the 26-pathogen legacy denominator. The 26-caliber 1990 opportunistic value is 22.846% at full precision (quoted as 22.9% at one decimal). effN = $1/\sum p^2$ over the 29 nodes.

#### Supplementary Table S5. Residual vaccine-preventable space scenario: fully expanded arithmetic, measured platform-sensitive scale, and ratios

| Step | Quantity | Computation | Result (deaths) |
|---|---|---|---|
| 1 | Pneumococcal deaths 2023, ages 0–19, global (measured, GBD 2023) | — | 227,976.5 |
| 2 | Hib deaths 2023, ages 0–19, global (measured, GBD 2023) | — | 17,774.8 |
| 3 | Global uncovered fractions, WUENIC 2023 | PCV3: 100 − 66 = 34% to 0.34; Hib3: 100 − 77 = 23% to 0.23 | — |
| 4 | Low scenario (residual pneumococcal space; smaller uncovered fraction as conservative bracket) | 227,976.5 × 0.23 | 52,434.6 (≈ 52,435) |
| 5 | High scenario (residual pneumococcal space; PCV3 uncovered fraction) | 227,976.5 × 0.34 | 77,512.0 (≈ 77,512) |
| 6 | Hib add-on | 17,774.8 × 0.23 | 4,088.2 (≈ 4,088) |
| 7 | Platform-sensitive deaths 2023, opportunistic/hospit al group, 5 pathogens (measured) | sum of GBD 2023 point estimates | 222,809.6 (≈ 222,810) |
| 8 | Platform-sensitive deaths 2023, full platform channel, 21 pathogens (measured) | sum of GBD 2023 point estimates | 396,994.5 (≈ 396,995) |

| Step | Quantity | Computation | Result (deaths) |
|---|---|---|---|
| 9 | Ratio, opportunistic group vs high scenario | 222,809.6 ÷ 77,512.0 | 2.87 |
| 10 | Ratio, full platform channel vs high scenario | 396,994.5 ÷ 77,512.0 | 5.12 |
| 11 | Ratio, opportunistic group vs low scenario | 222,809.6 ÷ 52,434.6 | 4.25 |

Note: Steps 4–6 are scenario estimates, not measurements; steps 1–2 and 7–8 are measured sums of GBD 2023 point estimates. Scenario assumptions: deaths distributed in proportion to coverage; vaccine fully effective within reach; indirect effects, serotype replacement, and the higher baseline risk of uncovered populations ignored. Scenario scope is restricted to pneumococcus and Hib; residual influenza and pertussis deaths (142,760 in 2023) are excluded. Both the restricted scope and the baseline-risk omission bias the residual downward, so the true residual vaccine-preventable space is most plausibly larger and the ratios (2.9–5.1×) are upper bounds. Ratios order magnitudes; they are not cost-effectiveness results.

**Supplementary Table S6. Effective number of pathogens (effN) at four time points, including the 2010 and 2019 intermediate anchors, and the channel × share-tier tabulation, 2023.**

S6a. effN and its components, 29-node spectrum (effN = 1/HHI; HHI = $\Sigma p^2$).

| Year | HHI ( $\Sigma p^2$) | effN (1/HHI) |
|---|---|---|
| 1990 | 0.1795 | 5.57 |
| 2010 | 0.1632 | 6.13 |
| 2019 | 0.1119 | 8.94 |
| 2023 | 0.1006 | 9.94 |

Note: The two intermediate anchor points (2010: effN 6.13; 2019: effN 8.94) document that diversification was monotonic and accelerated after 2010; the 2023 spectrum is spread across the equivalent of roughly ten equally sized pathogens against fewer than six in 1990. HHI values shown are the reciprocal of the tabulated effN at full precision.

S6b. Channel × share-tier tabulation, 2023 (deaths and SSA+SA concentration).

| Channel | Share tier | Pathogens (n) | Deaths 2023 | SSA+SA deaths | SSA+SA share (%) |
|---|---|---|---|---|---|
| Mixed (vaccine + platform) | High (>50k) | 1 | 87,764 | 76,463 | 87.1 |
| Mixed (vaccine + platform) | Mid (5k-50k) | 2 | 39,161 | 31,562 | 80.6 |
| Platform- sensitive | High (>50k) | 2 | 137,436 | 114,150 | 83.1 |
| Platform- sensitive | Mid (5k-50k) | 12 | 243,257 | 196,258 | 80.7 |
| Platform- sensitive | Low (<5k) | 7 | 16,301 | 12,855 | 78.9 |
| Vaccine- reachable | High (>50k) | 3 | 393,829 | 308,226 | 78.3 |
| Vaccine- reachable | Mid (5k-50k) | 2 | 47,581 | 38,489 | 80.9 |

Note: Tiers: high > 50,000 deaths in 2023; medium 5,000–50,000; low < 5,000. Tier figures are rounded; channel totals are computed at full precision, so tier sums may differ from channel totals by up to one death. Channel assignments as in Supplementary Table S1.

**Supplementary Table S7. Dose–response analysis, full results: super-region vaccine coverage (WUENIC 2015) versus 2010 to 2023 change in target-pathogen share (29-node caliber, ages 0– 19)**

| Vaccine | GBD super-region | Coverage 2015, median all countries (%) | Coverage 2015, mean introd uced (%) | Coverage 2015, weighted mean (%) | Countries (n) | Introduced 2015 (n) | Share 2010 (%) | Share 2023 (%) | Δ share (pp) |
|---|---|---|---|---|---|---|---|---|---|
| PCV3 | Central Europe , Easter n Europe , and Central Asia | 0.0 | 72.1 | 12.2 | 29 | 11 | 43.1 | 22.6 | -20.58 |
| PCV3 | High- income | 91.0 | 90.4 | 85.6 | 35 | 28 | 29.9 | 13.7 | -16.24 |
| PCV3 | Latin Americ a and Caribb ean | 83.5 | 84.8 | 82.8 | 30 | 19 | 36.1 | 22.6 | -13.44 |
| PCV3 | North Africa and Middle East | 71.0 | 89.9 | 41.9 | 21 | 13 | 37.5 | 24.6 | -12.90 |
| PCV3 | South Asia | 5.0 | 44.3 | 14.5 | 5 | 3 | 35.9 | 27.0 | -8.82 |
| PCV3 | Southe ast Asia, East Asia, and Oceani a | 0.0 | 66.7 | 1.9 | 29 | 11 | 35.7 | 21.1 | -14.51 |
| PCV3 | Sub- Sahara n Africa | 71.5 | 72.8 | 61.6 | 46 | 37 | 34.3 | 22.6 | -11.69 |
| Hib3 | Central Europe , Easter n Europe , and Central Asia | 93.0 | 87.7 | 52.7 | 29 | 28 | 2.3 | 2.7 | 0.36 |
| Hib3 | High- income | 96.0 | 95.3 | 94.7 | 35 | 35 | 2.2 | 2.4 | 0.14 |
| Hib3 | Latin Americ a and Caribb ean | 92.0 | 89.7 | 90.1 | 30 | 30 | 1.7 | 2.2 | 0.49 |
| Hib3 | North Africa and Middle East | 98.0 | 90.1 | 89.2 | 21 | 21 | 2.0 | 2.0 | -0.00 |
| Hib3 | South Asia | 91.0 | 81.0 | 54.0 | 5 | 5 | 2.0 | 2.2 | 0.29 |
| Hib3 | Southe ast Asia, East Asia, and Oceani a | 89.0 | 87.4 | 26.0 | 29 | 27 | 1.9 | 1.4 | -0.45 |
| Hib3 | Sub- Sahara n Africa | 84.0 | 77.6 | 70.0 | 46 | 46 | 1.4 | 1.7 | 0.24 |

Note: Coverage is the 2015 WUENIC estimate (midpoint of the transition window) aggregated to GBD super-regions; a WUENIC value of 0 denotes non-introduction, not measured zero coverage. Share is the target pathogen's share of the 29-node super-region total (pneumococcal share for PCV3; Hib share for Hib3). Spearman rank correlations across the seven super-regions (coverage 2015 median, all countries, versus 2010 to 2023 share change): PCV3 ρ = 0.108, p = 0.818, n = 7; Hib3 ρ = −0.036, p = 0.939, n = 7. Pneumococcal share fell in all seven super- regions; what is absent is a linear dose relationship between the regional coverage gradient and the magnitude of share decline. The analysis is ecological (n = 7) and is reported as descriptive, not causal, evidence.

#### Supplementary Table S8. Country-level composition of the three intervention channels, 2023:

channel deaths, top-5 countries by deaths, and sub-Saharan Africa plus South Asia (SSA+SA) share (26 rei-level LRI aetiologies, ages 0–19).

| Channel | Deaths 2023 | Top-5 countries by deaths (deaths) | SSA+SA share (%) |
|---|---|---|---|
| Vaccine-reachable (n=3) | 275,363 | India (58,746); Nigeria (49,878); DR Congo (12,645); Pakistan (11,704); Niger (9,018) | 81.6 |
| Mixed (vaccine + platform) (n=2) | 39,138 | Nigeria (6,518); India (6,163); Pakistan (2,289); DR Congo (2,093); Niger (1,735) | 80.6 |
| Platform-sensitive (n=21) | 396,727 | India (78,284); Nigeria (60,085); Pakistan (22,141); DR Congo (15,439); Niger (14,438) | 81.5 |

Note: Deaths are GBD 2023 point estimates, ages 0–19 years, summed across the 204 countries; country-level sums reconcile with global modelled values within 0.087%. Channel n counts aetiologies within the 26 rei-level LRI etiologies covered by the country extract; the manuscript 29-node caliber additionally includes Whooping cough (vaccine-reachable), Tuberculosis (mixed) and COVID-19 (vaccine-reachable, 2023), which are not rei-level LRI aetiologies. SSA+SA share is the percentage of channel deaths occurring in the Sub-Saharan Africa and South Asia GBD super-regions. Full country × aetiology × year data are provided in Additional file 2 (03_Dataset.xlsx).
